# Human MAIT cells are devoid of alloreactive potential: prompting their use as universal cells for adoptive immune therapy

**DOI:** 10.1101/2021.04.29.21256184

**Authors:** Marie Tourret, Nana Talvard-Balland, Marion Lambert, Ghada Ben Youssef, Mathieu F. Chevalier, Armelle Bohineust, Thomas Yvorra, Florence Morin, Saba Azarnoush, Olivier Lantz, Jean-Hugues Dalle, Sophie Caillat-Zucman

**Author notes:** **Correspondence to:** Sophie Caillat-Zucman, Laboratoire d’Immunologie, Hôpital Saint-Louis, 1 Avenue Claude Vellefaux, 75010, Paris, France. M.T, N.T-B and M.L contributed equally to this work. **Declarations**. Ethics approval and consent to participate* The study was carried out with the approval of the Robert Debré, Hospital Ethics Committee (HREC 2013/49) and the CPP Ile de France IV (2015/03NICB), in agreement with the principles of the Declaration of Helsinki and French legislation. All subjects (or their parents for the children cohort) provided written informed consent. The study was registered in a public trial registry: ClinicalTrials.gov number NCT02403089. Mice were used after approval of all procedures by the Institutional animal Care and Use Ethics Committee (CE121#16624). Availability of data:* Data are available on reasonable request to. Competing interests:* none declared. Author contributions and consent for publication* M. Tourret, N Talvard-Balland, M. Lambert, G. Ben Youssef, M. Chevalier, A. Bohineust, F Morin and S. Azarnoush designed research studies, conducted experiments, analyzed data, and/or provided patient samples. T. Yvorra produced synthetic 5-OP-RU. O Lantz and J-H. Dalle analyzed data and wrote the manuscript. S. Caillat-Zucman designed research studies, supervised the study, analyzed data, and wrote the manuscript with the help of other coauthors. All authors provided input, edited and approved the final version of the manuscript.

## Abstract

**Background:** Mucosal associated invariant T (MAIT) cells are semi-invariant T cells that recognize microbial antigens presented by the highly conserved MR1 molecule. MAIT cells are predominantly localized in the liver and barrier tissues and are potent effectors of anti - microbial defense. MAIT cells are very few at birth and accumulate gradually over a period of about 6 years during infancy. The cytotoxic potential of MAIT cells, as well as their newly described regulatory and tissue repair functions, open the possibility of exploiting their properties in adoptive therapy. A prerequisite for their use as “universal” cells would be a lack of alloreactive potential, which remains to be demonstrated.

**Methods:** We used *ex vivo, in vitro* and *in vivo* models to determine if human MAIT cells contribute to allogeneic responses.

**Results:** We show that recovery of MAIT cells after allogeneic hematopoietic stem cell transplantation recapitulates their slow physiological expansion in early childhood, independent of recovery of conventional T cells. *In vitro*, signals provided by allogeneic cells and cytokines do not induce sustained MAIT cell proliferation. *In vivo*, human MAIT cells do not expand nor accumulate in tissues in a model of T-cell mediated xenogeneic graft-versus-host disease (GVHD) in immunodeficient mice.

**Conclusions:** Altogether, these results provide evidence that MAIT cells are devoid of alloreactive potential and pave the way for harnessing their translational potential in universal adoptive therapy overcoming barriers of HLA disparity.

## Introduction

MAIT cells are innate-like T cells expressing a semi-invariant TCR which recognizes microbial-derived riboflavin (vitamin B2) precursors such as 5-OP-RU presented by MR1, a monomorphic non-classical MHC class-1 like molecule (1-3). Upon TCR engagement, MAIT cells proliferate, release proinflammatory cytokines and kill target cells, supporting their role in antimicrobial defense (4-6). Most bacteria and yeasts, but not mammal cells, are able to synthesize riboflavin and hence provide MR1 ligands (7). This TCR-MR1 recognition pathway therefore represents a sophisticated discriminatory mechanism to target microbial antigens while sparing the host. MAIT cells can also be activated in a TCR-independent way in response to inflammatory cytokines such as IL-12 and IL-18 (8-10). MAIT cells are predominantly localized in the liver and barrier tissues, in agreement with their expression of several chemokine receptors (4, 11). They are also abundant in the adult blood, but very few in cord blood. We previously showed that the postnatal expansion of MAIT cells was a very slow process requiring at least 5-6 years to reach adult levels, and likely resulted from repeated encounters of a few MAIT cell clones with MR1-restricted microbial antigens (12).

The curative effect of allogeneic hematopoietic stem cell transplantation (HSCT) in hematological malignancies is based on the capacity of donor T cells to eliminate residual tumor cells (graft-versus-leukemia (GVL) effect). A serious drawback is that donor T cells may also recognize normal nonhematopoietic cells, leading to graft-versus-host disease (GVHD), which is characterized by activation, expansion and migration to target tissues of donor alloreactive T cells (13-17). In the first weeks after HSCT, the T cell compartment recovers through peripheral expansion of graft-derived T cells in response to increased levels of homeostatic cytokines and to host’s allogeneic antigens. Reconstitution of a fully diversified T-cell repertoire occurs only later by resumed thymic output of newly-generated naïve T cells (18). Even under favorable conditions, it takes at least 2 months to produce naive T cells, and a plateau of thymic output is reached only after 1-2 years.

We observed that MAIT cell recover y was delayed up to 6 years after allogeneic cord blood transplantation, mimicking the long postnatal expansion period (12). Other studies confirmed that MAIT cell numbers failed to normalize for at least a year after HSCT (19-21) and suggested that a low MAIT cell number was associated with an increased risk of GVHD, although the mechanisms remain unclear (19, 21, 22). In mouse HSCT models, recipient’s residual MAIT cells protect ed mice from acute intestinal GVHD through microbial-induced secretion of IL-17 promoting gastrointestinal tract integrity and reducing alloantigen presentation, ultimately limiting the proliferation of donor-derived alloreactive T cells (23). Recently, transcriptomic and functional studies have revealed new tissue repair and regulatory functions of MAIT cells (24-28). Altogether, these results open the possibility of exploiting the effector or tissue repair /regulatory properties of MAIT cells in adoptive immunotherapy, provided that they do not cause alloreactivity. Here, we used *in vitro* and *in vivo* models to provide evidence that human MAIT cells are devoid of alloreactive potential. Our results pave the way for harnessing the translational potential of MAIT cells in universal adoptive therapy overcoming barriers of HLA disparity.

## Patients and Methods

### Patients

Three independent cohorts of patients undergoing allogeneic HSCT for hematological malignancy were studied. Patient and graft characteristics are described in Table 1.

**Table 1.** Characteristics of HSCT recipients. ALL: acute lymphoblastic leukemia; AML: acute myeloid leukemia; CLL: chroniclymphoblastic leukemia; CML: chronic myeloid leukemia; JMML: juvenile myelomonocyticleukemia ; MDS : myelodysplasic syndrome ; NHL : non-Hodgkin lymphoma. MA : myeloablative ; NMA : non-myeloablative.

|  | **Children**<br>**Longitudinal**<br>**cohort** | **Children**<br>**Cross-sectional**<br>**cohort** | **Adults**<br>**Retrospective**<br>**cohort** |
| --- | --- | --- | --- |
| **Patients (n)** | 40 | 64 | 49 |
| **Sex, n (%)** |  |  |  |
| - **Female** | 17 (42.5) | 23 (36) | 18 (36.7) |
| - **Male** | 20 (57.5) | 41 (64) | 31 (63.3) |
| **Age, mean years (range)** | 9.4 (0.75-16) | 7 (0.9-17.8) | 44.4 (17-65.8) |
| **Primary disease, n (%)** |  |  |  |
| - **ALL** | 23 (57.5) | 25 (39.1) | 6 (12.2) |
| - **AML** | 11 (27.5) | 11 (17.2) | 19 (38.8) |
| - **JMML** | 3 (7.5) | 2 (3.1) | - |
| - **NHL** | 1 (2.5) | 1 (1.6) | 13 (26.5) |
| - **MDS** | 1 (2.5) | 2 (3.1) | 5 (10.2) |
| - **CML** | 1 (2.5) | 3 (4.7) | 2 (4.1) |
| - **Others** | - | 20 (31.2) | 4 (8.2) |
| **Donor, n (%)** |  |  |  |
| - **MSD** | 19 (47.5) | 32 (50) | 49 (100%) |
| - **MUD** | 21 (52.5) | 32 (50) | - |
| **Stem cell sources, n (%)** |  |  |  |
| - **Peripheral blood** | - | - | 30 (61.2) |
| - **Bone marrow** | 40 (100) | 64 (100) | 19 (38.8) |
| **Conditioning, n (%)** |  |  |  |
| - **MA** | 40 (100) | 64 (100) | 24 (49) |
| - **NMA** | - | - | 25 (51) |
| **In vivo T-cell depletion, n (%)** | 12 (30) | 18 (28) | 20 (40.8) |
| **Acute GVHD** |  |  |  |
| - **Grade 1-2** | 24 (60) | NA | 21 (42.8) |
| - **Grade 3-4** | 9 (22.5) | NA | 4 (8.2) |

Cohort 1 included 40 children recipients, for whom peripheral blood mononuclear cells (PBMCs) samples were prospectively collected at Robert Debré Hospital between January 2013 and December 2015. Blood samples were collected prior to conditioning (± day −15 before HSCT) and at 1, 3, 6, 12 and 24 months after HSCT as the standard of care for assessment of immunologic recovery. Patients who died before day 180 were not included in the analysis.

Cohort 2 included 64 additional HSCT children in stable remission for whom PBMCs samples were collected at time of a routine visit at Robert Debré Hospital 2 to 16 years after HSCT. All children from cohorts 1 and 2 received unmanipulated bone marrow transplant from HLA-matched sibling donor (MSD) or unrelated donor (MUD). Myeloablative conditioning was provided with VP16 and total body irradiation (TBI), or with cyclophosphamide and busulfan. *In vivo* T-cell depletion by ATG was given in the majority of MUD recipients. Primary prophylaxis of GVHD consisted of a calcineurin inhibitor alone (MSD recipients) or with methotrexate (MUD recipients).

Cohort 3 included 49 adult donor/recipient pairs for whom frozen annotated PBMCs were provided by the CRYOSTEM consortium (https://doi.org/10.25718/cryostem-collection/2018) and SFGM-TC (Société Francophone de Greffe de Moelle-Thérapie Cellulaire). Patients received bone marrow or peripheral blood stem cells from a matched sibling donor and were given myeloablative or non-myeloablative conditioning. Blood samples were collected before conditioning, and at 3 and 12 months post-HSCT in the absence of aGVHD. In case of aGVHD, samples were collected at the time of diagnosis before any treatment, one month later (i.e towards 3 months post-HSCT) and at 12 months.

### Cells and reagents

PBMCs were isolated and used immediately or frozen. 5-OP-RU was synthesized as described in (29-31). Human MAIT cells were expanded for 6 days in human T-cell culture medium (RPMI-1640, Invitrogen, Life Technologies) containing 10% human AB serum (EuroBio), IL-2 (100 U/mL, Miltenyi) and 300 nM 5-OP-RU.

### Flow cytometry

MAIT cells were analyzed in fresh whole blood, or in isolated PBMCs where indicated. Multiparametric 14-color flow cytometry analyzes were performed as described in Supplementary data. MAIT cells were defined as CD3 ^+^CD4^-^CD161^high^Vα7.2^+^ cells in the first part of the study (HSCT patients). This population fully overlapped with the population labeled by MR1:5-OP-RU tetramers (31). Thereafter, we used the specific MR1:5-OP-RU tetramer when it became available (NIH tetramer core facility).

### In vitro stimulations

Human carboxyfluorescein succinimidyl ester (CFSE)-labeled (5μM) PBMCs were cultured in RPMI-1640 supplemented with IL-2 (100 U/mL), IL-15 (50 ng/mL), IL-7 (50 ng/mL, all from Miltenyi Biotec), or IL-12/IL-18 (50 ng/mL each, R&D Systems) and/or 300 nM 5-OP-RU. For mixed lymphocyte reactions, CFSE-labeled PBMCs used as responders (1×10^6^/ml) were incubated with γ-irradiated allogeneic stimulator PBMCs (1:1 ratio) in 96-well round-bottom plates. Cells were harvested at day 6 and stained before flow cytometry analysis.

### Adoptive transfer of xenogeneic cells

NOD-Scid-IL-2Rγ^null^ (NSG) mice (Jackson laboratory, Bar Harbor, MI) were bred in the animal facility of St-Louis Research Institute and housed under specific pathogen-free conditions. Eight- to 10-week-old female mice were used after approval of all procedures by the Institutional animal Care and Use Ethics Committee (CE121#16624). Mice were irradiated (1.3 Gy) 24 hours prior to injection of 5×10^6^ human PBMCs in the caudal vein. Development of GVHD was monitored 3 times per week based on weight loss, hunching posture, reduced mobility and hair loss. Human chimerism in peripheral blood (percent of human CD45 ^+^ cells) was assessed weekly. Where indicated, mice received 5-OP-RU (1 nmol i.p. every 3 days from the day of PBMCs infusion) and/or human IL-15 (100 ng i.p. every 3 days). Mice were sacrificed at the indicated time, or when weight loss was >15%. Peripheral blood, spleen, liver, lungs and intestine were harvested, and cells were isolated as described in Supplementary data.

### Statistics

Differences between groups were analyzed using non-parametric tests for paired (Wilcoxon) and unpaired (Mann-Whitney or Kruskall-Wallis) groups, or two-way ANOVA. Correlations were assessed using the Spearman’s rank correlation. Two-sided p values < 0.05 were considered significant. Analyzes were performed using Prism software v. 9 (GraphPad). All data including outliers were included with one pre-determined exception: flow cytometry cell-subset percentages were considered non-evaluable if the parent subset contained <100 events.

## Results

### 1/ MAIT cell reconstitution is delayed for several years after HSCT

Our previous findings have shown that it takes up to 6 years to recover normal MAIT cell values after cord blood transplantation, suggesting that MAIT cells do not proliferate in response to allogeneic stimulation (12). However, MAIT cells in cord blood are naïve, and their number is 1-2 logs lower than in adult blood, which could contribute to this slow recovery. We therefore analyzed the kinetics of MAIT cell reconstitution in other HSCT settings.

Forty children who received an unmanipulated bone marrow transplant after myeloablative conditioning were studied longitudinally up to 24 months after HSCT. While the number of conventional T cells (Tconv) gradually increased from 1 month after transplantation and returned almost to normal after one year, there was virtually no increase in the number of MAIT cells during the study period (Figure 1A). Two years after HSCT, MAIT cell values remained 5 times lower than in age-matched donors (Figure 1A). The number of Tconv and MAIT cells before and up to 3 months after HSCT was lower in matched unrelated donor (MUD) than in matched sibling donor (MSD) recipients, likely due to a longer time to transplant and more frequent use of *in vivo* T-cell depletion in MUD recipients. However, while Tconv reached comparable values after 3 months regardless of the donor type, MAIT cell numbers remained around 2 times lower in MUD recipients than in MSD recipients during the 24-months follow-up (Figure 1B).

**Figure 1:**
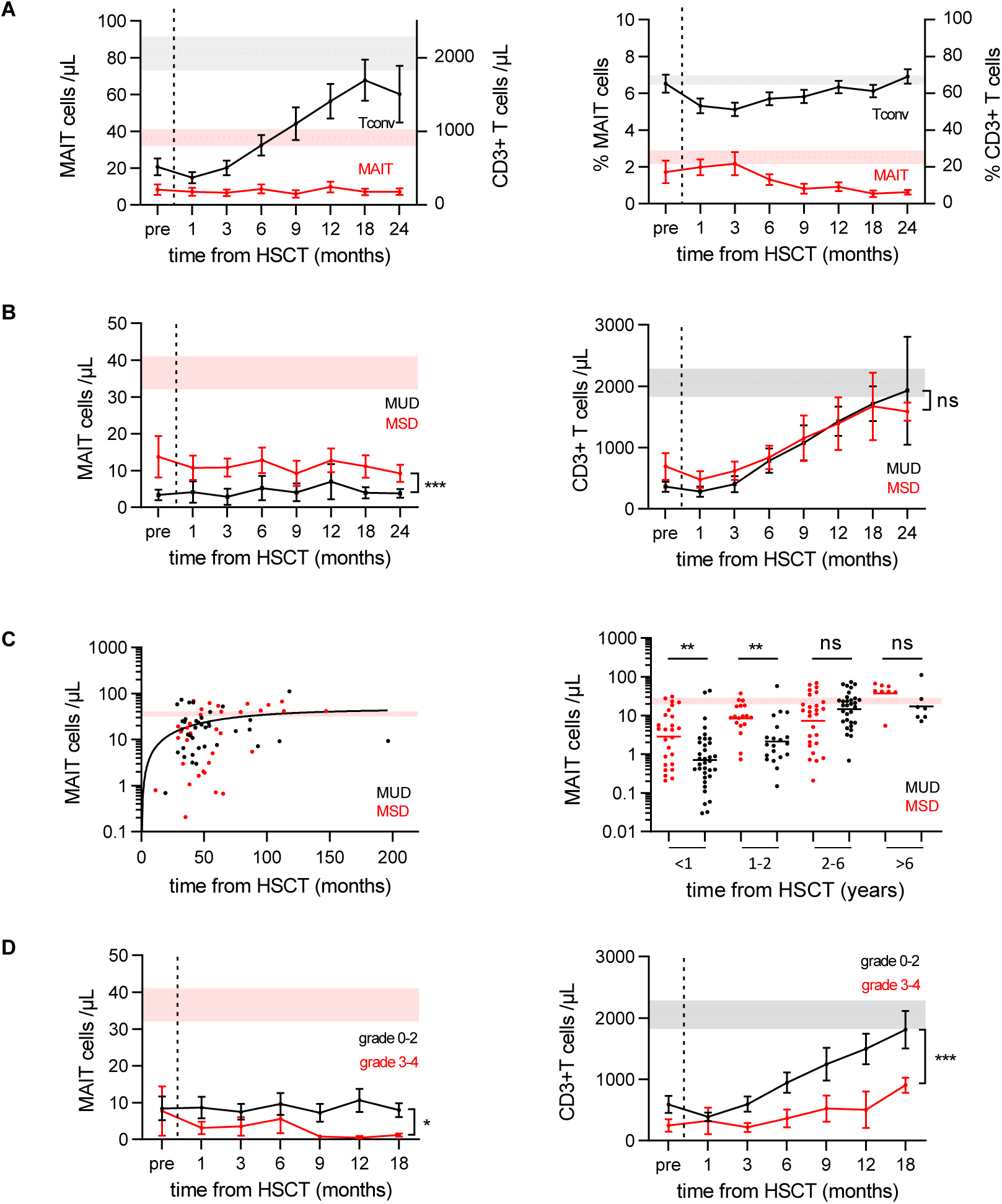
MAIT cell re covery takes several years after HSCT in children. (A)Absolute counts (left panel) and percentages (right panel) of CD3 T cells (Tconv) and MAIT cells detected in the peripheral blood of 4 0 children before conditioning (pre) and from month 1 to month 24 after HSCT. Data are shown as mean ± SEM. (B)Recovery dynamics of MAIT cells (left panel) and CD3 T cells (right panel) in transplant recipients of matched sibling donors (MSD, red line) and unrelated donors (MUD, black line). Results show the mean absolute numbers ± SEM during the first 24 months after HSCT. 2-way ANOVA, *** P = 0.0005; ns: not significant. (C)MAIT cell recovery takes at least 6 years to reach normal values. Left panel: Relationship between log10-transformed MAIT cell absolute numbers and time from transplantation up to 16 years after HSCT in MSD (red dots) and MUD (black dots) recipients. Right panel: Recovery is slower in MUD than MSD recipients for at least to 2 years after HSCT. Mann-Whitney ** P ≤ 0.002; ns: not significant (D)Comparison of MAIT (left panel) and Tconv (right panel) cell recovery in patients without or with mild (grade 0-2) or severe (grade 3-4) aGVHD. Results show the mean absolute numbers ± SEM. 2-way ANOVA, * P= 0.04, ***P = 0.000 6. Light gray and red areas show the reference physiological T cell and MAIT cell count interval, respectively, obtained from 36 age-matched healthy donors.

To extend our findings to a longer follow-up period, we analyzed 64 additional children at time of a routine visit 2 to 16 years after transplantation. As observed after cord blood transplantation (12), the number of MAIT cells increased very slowly (even more slowly in MUD than MSD recipients), to reach a plateau approximately 6 years after HSCT (Figure 1C). This slow recovery was not related to the underlying malignancy, gender or age of the recipient, pre-HSCT conditioning (with or without TBI), severe microbial infection during the first months after HSCT, or duration of immunosuppressive treatment (data not shown).

T-cell reconstitution is impaired in patients with aGVHD, at least in part because of defective thymic production of HSC-derived T cells (18). We previously observed that thymus-derived naïve MAIT cells appeared in cord blood recipients 6 months after transplantation (12). Here we found that the number of MAIT cells after 6 months was lower in patients with severe (grade 3-4) aGVHD compared to those without or mild (grade 0-2) aGVHD, as also observed for Tconv cells (Figure 1D). These data suggest that the alteration of thymic function associated with aGVHD participates in the defective recovery of MAIT cells after HSCT.

### 2/ The reconstitution of MAIT cells after HSCT is affected by their number in the donor and by the existence of aGVHD

To further explore the potential link between MAIT cell numbers aGVHD and MAIT cell numbers, we retrospectively analyzed 49 adult HSCT donor/recipient pairs from the national Cryostem consortium, including 25 patients with aGVHD. As observed in children, MAIT cells did not significantly expand during the one-year study period, while the number of Tconv increased sharply. Moreover, absolute numbers of MAIT and Tconv cells at 3 months were lower in patients with severe aGVHD than in those without or mild aGVHD, whereas subsequent reconstitution was not affected (Figure 2A). The proportion of Ki67^+^ cells at the time of aGVHD, which could represent alloreactive proliferating cells, was higher in Tconv cells than in MAIT cells. One year after HSCT, Ki67 expression was hardly detectable in both cell types (Figure 2B).

**Figure 2:**
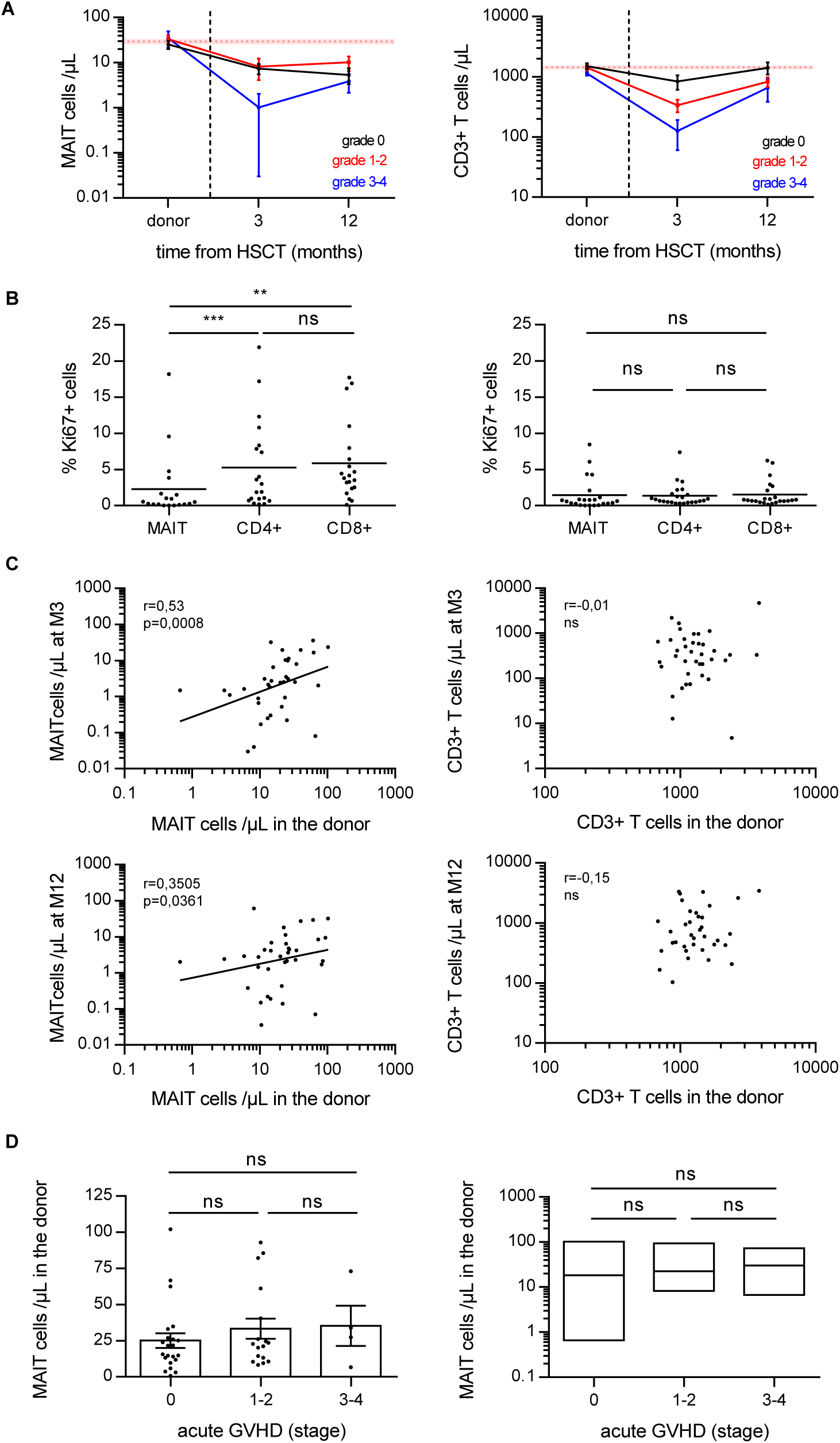
MAIT cell recovery in adult HSCT recipients. (A)Kinetics of MAIT (left panel) and Tconv (right panel) cell recovery in patients from the CRYOSTEM cohort (n= 49) according to the absence or the presence of mild (grade 1-2) or severe (grade 3-4) aGVHD. Patients with aGVHD were sampled at the time of disease diagnosis and 1 month later, i.e. towards M3. Results show the mean absolute numbers ± SEM. Shaded areas show MAIT and T cell count intervals in the donors. 2-way ANOVA, not significant. (B)Correlation between the number of MAIT (left) or Tconv (right) cells in the donor and that in the recipient 3 months (top panels) and 12 months (bottom panels) after HSCT. Spearman correlation r and P values are indicated. (C)Quantification of Ki67 ^+^ proliferating cells among MAIT, CD4 and CD8 T cells at time of aGVHD (left panel) and 12 months after HSCT (right panel). Scatter plots with bar showing the mean percentage of Ki67^+^ cells. Wilcoxon non parametric paired test, ** P= 0.004; *** P= 0.0008; ns: not significant. (D)Comparison of the number of MAIT cells in the do nor according to the absence or presence of mild (grade 1-2) or severe (grade 3-4) aGVHD. Kruskall-Wallis non-parametric paired test, ns: not significant

MAIT cell recovery was not influenced by the conditioning regimen (myeloablative or non-myeloablative) or by the source of HSC (peripheral blood or bone marrow) (data not shown). Notably, the number of MAIT cells in the donor was significantly correlated to their number in the recipient after HSCT, whereas no correlation was observed for Tconv (Figure 2C). However, there was no association between the number of MAIT cells in the donor and the presence of severe aGVHD (Figure 2D).

Altogether, these data indicate that the number of MAIT cells in the donor influences the early reconstitution of MAIT cells in the recipient, but do not provide evidence for a role of MAIT cells in aGVHD.

### 3/ MAIT cel ls do not proliferate in response to alloantigen stimulation, except in the presence of MR1 ligand

Graft-derived conventional T cells expand through lymphopenia-induced homeostatic proliferation and response to host’s allogeneic antigens. However, not all T cells respond with the same efficiency to these proliferative cues (32).

We first analyzed the *in vitro* responsiveness of MAIT cells to homeostatic cytokines. CFSE-labeled PBMCs were treated with IL-7 or IL-15, or IL-2 as control, alone or in combination with the microbial-derived MR1 ligand, 5-OP-RU, and the proliferation of MR1-tetramer^+^ MAIT cells was monitored according to CFSE dilution (Figure 3A, B). In the absence of cytokine, MAIT cells did not respond to 5-OP-RU alone. Most MAIT cells proliferated in response to IL-15 or IL-7 (but not to IL-2) alone, but the number of divisions was significantly higher when 5-OP-RU was added to IL-15 or IL-2.

**Figure 3:**
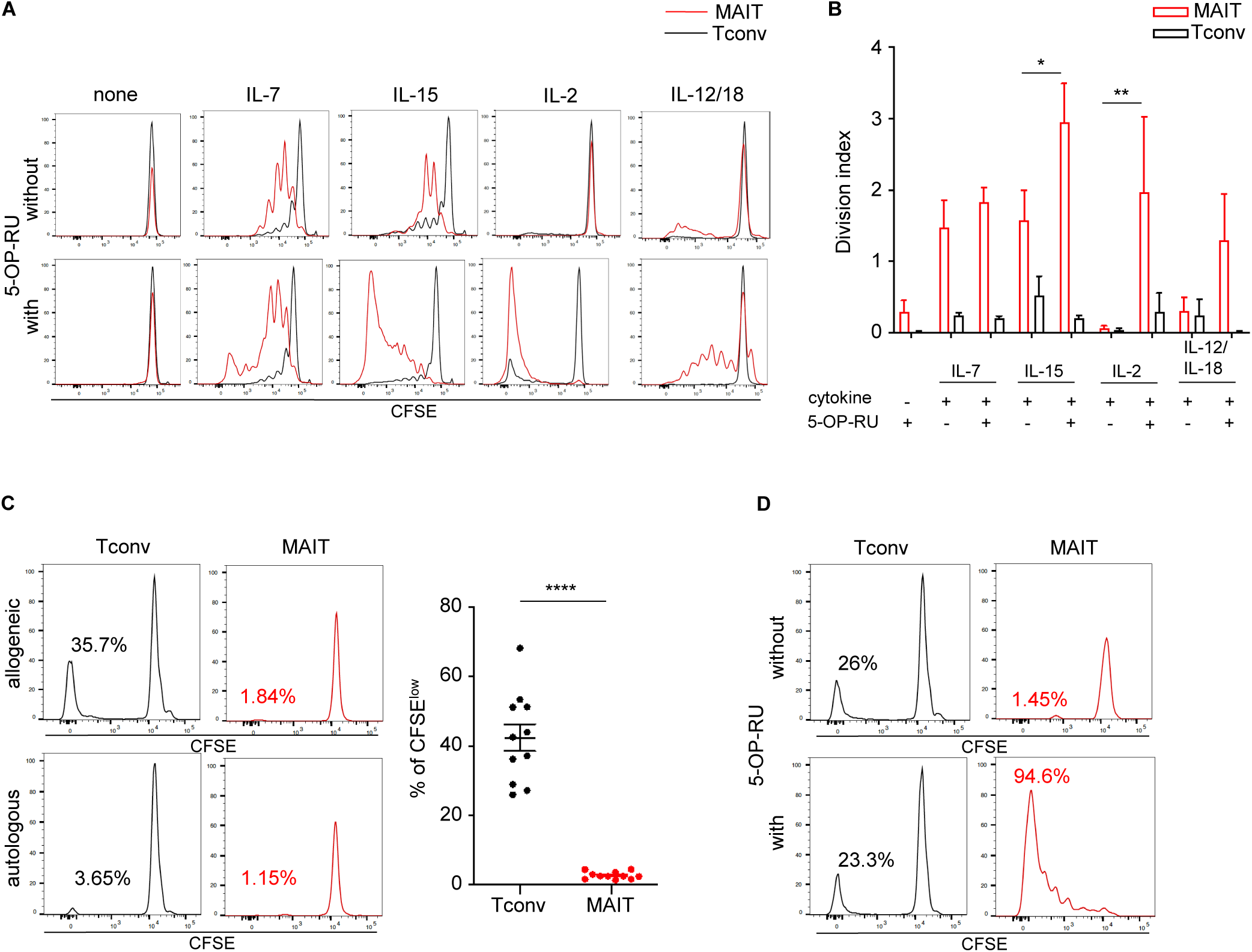
MAIT cell prolifer ative response to cytokines and allogeneic cells *in vitro*. Carboxyfluorescein succinimidyl ester (CFSE)-labeled human PBMCs (10^6^/mL) were cultured for 6 days in the indicated conditions. Proliferation of MR1 tetramer ^+^ MAIT (red lines) and conventional T (Tconv, black lines) cells was quantified by CFSE dilution at the end of the culture. (A, B) Proliferative response to stimulation by IL-7 (50ng/mL), IL-15 (50 ng/mL), IL-2 (100 U/mL) and IL-12/18 (50 ng/mL each) in the presence or absence of the synthetic MR1 ligand, 5-OP-RU (300 nM). (A) Representative CFSE staining gated on MA IT and Tconv cells after a 6-day culture. (B) Division index (mean number of divisions in the total population) of MAIT and Tconv cells. Mean ± SEM of 3 independent experiments. Paired t test, * P= 0.045; ** P= 0.004. (C) Mixed lymphocyte reaction: CFSE-labeled responder PBMCs were cultured with irradiated allogeneic PBMCs at 1:1 ratio. Responder T cells were identified at the end of the 6-day culture by gating on CD3^+^ T cells following exclusion of dead cells. Left panel: Representative CFSE staining gated on MAIT (red) and Tconv (black) cells after 6-day culture in the presence of allogeneic (upper panel) or autologous (lower panel) PBM Cs. Numbers in the quadrants indicate the percentage of proliferating (CFSE ^low^) cells among each population. Right panel: Individual values and means ± SEM of proliferating Tconv and MAIT cells (n= 11 experiments using different recipient/donor pairs). Paired t test, *** * P<0.0001 D) Mixed lymphocyte reaction was performed in the absence (upper panel) or presence (lower panel) of 300 nM 5-OP-RU. Representative CFSE staining gated on MAIT (red) and Tconv (black) cells after a 6-day culture.

Chemotherapy and radiation induce the secretion of IL-12 and IL-18, which may trigger early TCR-independent activation of MAIT cells transferred with the graft. We observed a low proliferati ve response of MAIT cells to IL-12/IL-18 combination, which increased in the presence of 5-OP-RU but remained lower than proliferation induced by IL-15 (Figure 3A, B). We next explored the capacity of MAIT cells to respond to allogeneic cells in a mixed lymphocyte reaction (MLR). Unlike Tconv, MAIT cells barely proliferated in response to allogeneic stimulation (Figure 3C). However, the addition of 5-OP-RU to the MLR induced a strong proliferation of MAIT cells (Figure 3D), suggesting that cytokines (IL-2 or other) produced by neighboring alloreactive T cells during the culture period allowed MAIT cells to proliferate only in response to the MR1 ligand.

Overall, these results suggest that signals provided by allogeneic cells and cytokines present in the post-HSCT period are not sufficient to induce sustained expansion of MAIT cells.

### 4/ Human MAIT cells do not expand in immunodeficient mice and do not cause xenogeneic GVHD (xeno-GVHD)

To further explore the potential of human MAIT cells to expand *in vivo* and participate to GVHD tissue lesions, we used a model of xenogeneic GVHD (xeno-GVHD) in which low doses of human PBMCs (huPBMCs) are injected into irradiated immunodeficient NSG mice. In this model, human T cells consistently expand in the mouse and mediate an acute GVHD-like syndrome with extensive T-cell tissue infiltration and damage of mouse skin, liver, intestine and lungs, resulting in death by 30-50 days (33, 34).

Mice were injected with 5×10^6^ huPBMCs, among which MAIT cells represented around 3% of T cells. The presence of CD45^+^ huPBMCs was determined at different times in tissues of recipient mice, including those where MAIT cells are known to preferentially reside. Four weeks after transfer, variable proportions of CD45^+^ cells were found in peripheral blood and tissues, almost all of which were Tconv and less than 0.05% were MAIT cells (Figure 4A).

**Figure 4:**
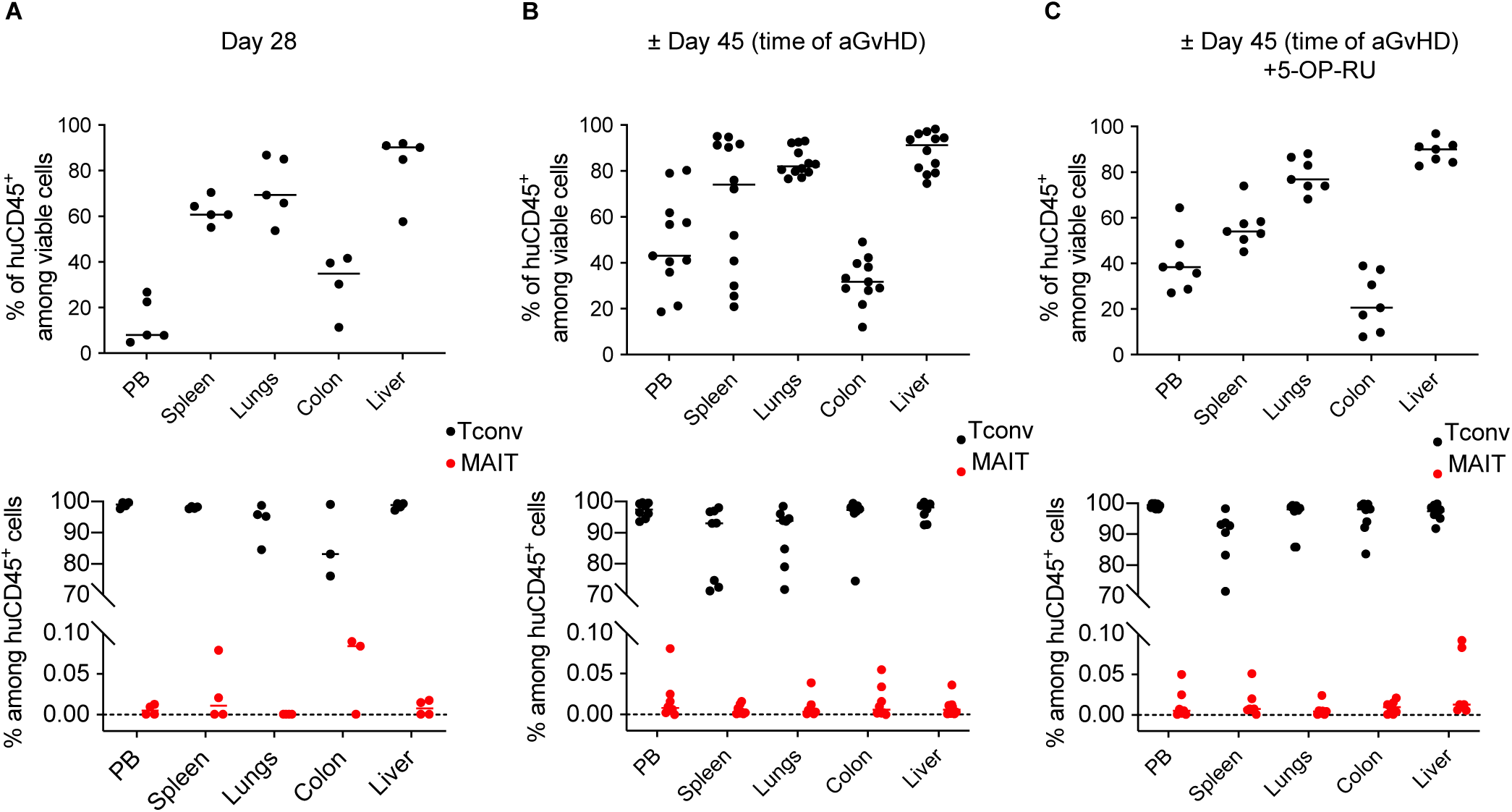
Absence of MAIT cell expansion in a xenogeneic GVHD model. Irradiated (1.3 Gy) NSG mice injected with 5.10^6^ huPBMCs were monitored to evaluate aGVHD progression and euthanized at the indicated time. Peripheral blood, spleen, liver, lungs and colon were harvested, and cells were isolated. (A-C) The proportion of human CD45+ leukocytes among viable cells (i.e. human chimerism, upper panels), and the proportion of Tconv (black dots) and MAIT (red dots) cells among human CD45+ cells (lower panels) were determin ed by flow cytometry. Results show individual values and median (horizontal bar). (A) Mice were sacrificed at day 28 after injection (n= 5 mice). (B) Mice were euthanized when they exhibited signs of aGVHD (± day 45, n= 11 diseased mice). (C) Mice received 5-OP-RU every 3 days (1 nmol i.p.) from the day of huPBMC injection and were euthanized at time of aGVHD (n=7 diseased mice).

Next, mice injected with huPBMCs were monitored to evaluate aGVHD progression and euthanized when weight loss was >15% (±45 days after transfer). A massive accumulation of Tconv was observed in tissues, in particular in the spleen, lungs and liver. However, MAIT cells were very few in all compartments (Figure 4B).

Human MAIT cells efficiently recognize the murine MR1 molecule (35), ruling out a defective presentation of MR1 ligands to human MAIT cells in NSG mice. However, since MAIT cells do not proliferate significantly *in vitro* in the absence of 5-OP-RU, the availability of MR1 ligand could be a factor limiting their expansion or survival in mice housed under specific-pathogen-free conditions. Mice were thus injected with 1 nmol 5-OP-RU intraperitoneally every 3 days from the day of huPBMC injection, a dose previously shown to activate endogenous MAIT cells (36). This did not result in any increase in MAIT cells in peripheral blood or tissues (Figure 4C).

It is not clear whether mouse cytokines can sustain homeostatic proliferation and survival of human MAIT cells in NSG mice due to a species barrier between human lymphoid cells and the murine recipient microenvironment (37-39). This is a key question for IL-15, as it is mostly mouse-derived in the xeno-GVHD model given the low human myeloid chimerism. Indeed, we found that human MAIT cells cultured with mouse IL-15 did not proliferate at all *in vitro* (Figure 5A). Proliferation was increased when 5-OP-RU was added to mouse IL-15, although it remained lower than with human IL-15. Murine and human IL-7 had similar effects on MAIT cell proliferation, regardless of the presence of 5-OP-RU (Figure 5A). Thus, IL-15 availability may be a factor limiting the proliferation of MAIT cells in NSG mice.

**Figure 5:**
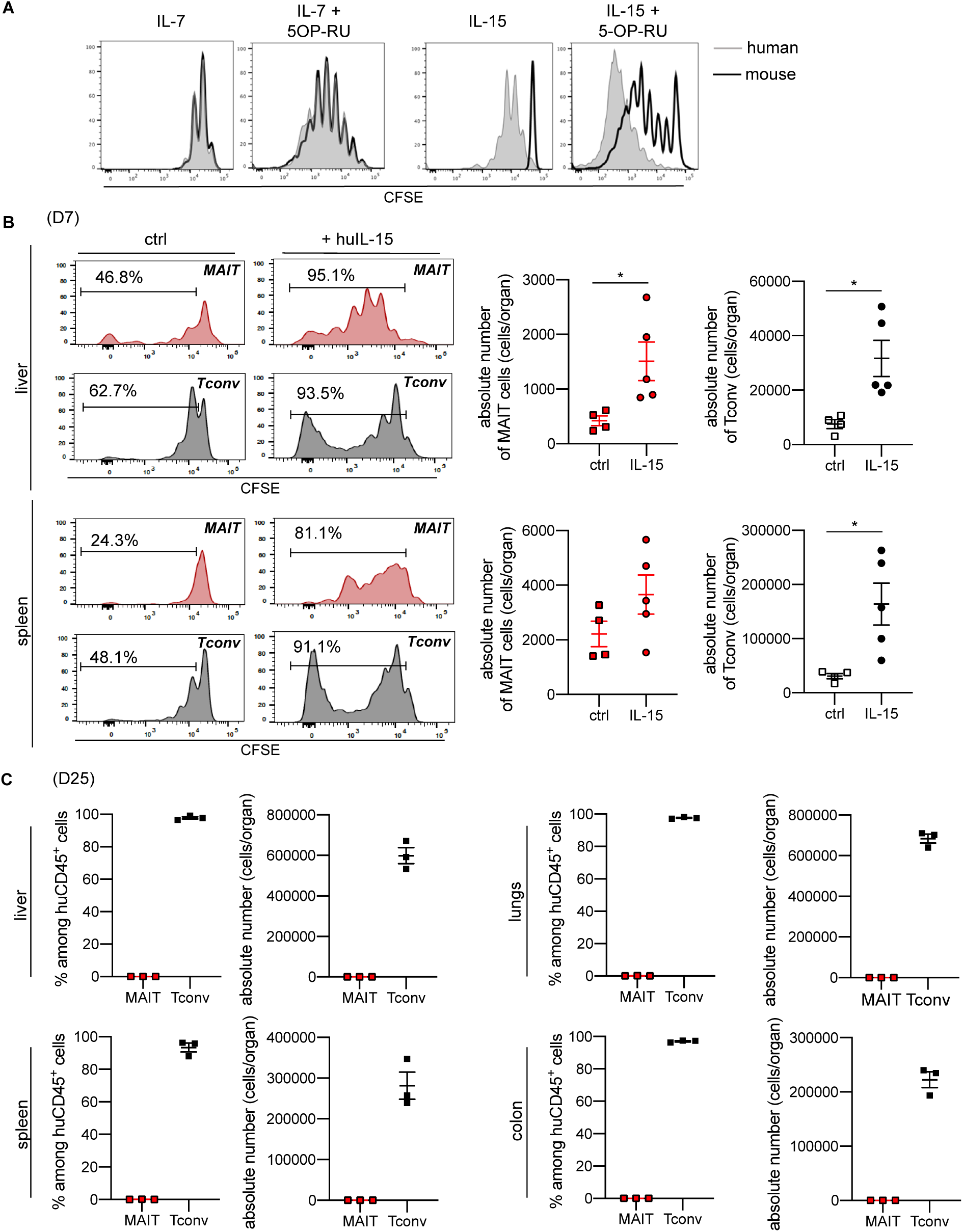
Human IL-15 does not increase MAIT cell accumulation in the target tissues in the xeno-GVHD model. (A)CFSE-labeled human PBMCs were cultured for 6 days with mouse (open black histograms) or human (shaded grey histograms) IL-15 (50 ng/mL) or IL-7 (10 ng/mL) in the absence or presence of 5-OP-RU. Representative experiment showing CFSE^low^ proliferating MAIT cells (gated on CD3 T cells). (B)NSG mice were injected with 20×10^6^ CFSE-labeled human PBMCs without or with human IL-15 (100 ng i.p. every other day from the day of transfer). The spleen and liver were harvested 7 days later, and cells were analyzed by flow cytometry for CFSE expression (left panel) and numbers of MAIT and Tconv cells (right panel). (C, D) Mice were treated with huIL-15 from the day of transfer of 5.10^6^ huPBMCs, and euthanized at the time of aGVHD (day 25, n= 3 diseased mice). Results show the proportion of human CD45+ leukocytes among viable cells (C), and absolute counts of Tconv and MAIT cells (D) in the indicated compartments. Individual values and median (horizontal bar) are shown.

We therefore sought to determine the division rate of MAIT cells after transfer of CFSE-labeled huPBMCs into NSG mice. One week after transfer, large proportions of both Tconv and MAIT cells from the liver and spleen displayed low CFSE fluorescence, indicating that they were able to divide (Figure 5B). These proportions increased when mice were treated with human IL-15 from the day of transfer, leading to a significant accumulation of Tconv and MAIT cells in the liver and spleen (figure 5B).

We, therefore, investigated whether human IL-15 would enhance MAIT cell accumulation in target tissues during xeno-GVHD. NSG mice were treated with human IL-15 three times per week from the day of huPBMC transfer and euthanized when they developed severe signs of aGVHD (25 days after transfer, i.e. earlier than control mice). A massive infiltration of T cells was observed in all compartments, but MAIT cells still remained barely detectable (Figure 5 C). Altogether, these results indicate that human MAIT cells are able to divide and survive in immunodeficient mice, but do not participate in T cell-mediated tissue lesions during xeno-GVHD even in the presence of human IL-15.

## Discussion

The newly described tissue repair and immunoregulatory functions of MAIT cells open fascinating perspectives for their use in adoptive therapy to control immune-mediated damage in tissues where these cells are known to accumulate. However, such strategy can be considered in unrelated recipients only if MAIT cells are devoid of GVH potential. As anticipated by the very limited diversity of their TCR that recognizes microbial antigens presented by the highly conserved MR1 molecule, our results demonstrate that MAIT cells do not proliferate in response to allogeneic signals and do not participate to aGVHD lesions, in line with our previous observation that MAIT cells were undetectable in target tissues at time of aGVHD (12).

Human MAIT cells are very few at birth and accumulate gradually during infancy, with about a 30-fold expansion to reach a plateau around 6 years of age (12). Several pieces of evidence suggest that the drivers of this peripheral expansion are related to successive encounters with microbes leading to an accumulation of MAIT cell clonotypes that will constitute the future MAIT cell pool (40). Indeed, MAIT cells are absent in germ-free mice and are very few in laboratory mice, but dramatically expand following challenge with riboflavin - producing microbes (35, 36, 41). Moreover, their development in mice depends on early-life exposure to defined microbes that synthesize riboflavin-derived antigens (27). In human subjects with controlled infection, MAIT cells show evidence of expansion of select MAIT cell clonotypes (42), but longitudinal analyzes of MAIT cell numbers in relation to the composition of the microbial environment are still lacking.

Here, we show that the reconstitution of MAIT cells after HSCT recapitulates their physiological expansion in the infancy and takes place over a period of at least 6 years, regardless of recipient- or donor-related factors such as age, underlying disease, donor type, stem cellsource or conditioning regimen. The number of MAIT cells in the early post-transplant period is at least in part dependent on the number of MAIT cells transferred with the graft. Subsequent MAIT cell recovery relies upon thymus-derived naïve MAIT cells. The low absolute MAIT cell counts in patients with grade 3-4 aGVHD, which associate with low absolute counts of conventional T cells, may be a consequence of a defective thymic function. Indeed, aGVHD-associated thymic damage results in the loss of the large population of double-positive (DP) thymocytes (43). This may decrease the thymic output of naïve MAIT cells, which undergo positive selection by recognizing MR1 at the surface of these DP thymocytes (44). In addition, loss of diversity and increased bacterial domination early after HSCT, in particular by Enterococcus, has been associated with increased risk of aGVHD (45-47). One might speculate that blooming of Enterococcus (a strain unable to synthesize riboflavin) could prevent from early expansion of donor-derived MAIT cells due to lack of MR1-ligands. So far, the relationship between abundance and diversity of gut microbiota and post-transplant MAIT cell reconstitution remains controversial (19, 22). Investigating the gut metagenome or metatranscriptome of HSCT recipients for the presence of riboflavin biosynthesis genes should answer this question.

In mouse HSCT models, conditioning-resistant host residual MAIT cells promote gastrointestinal tract integrity and limit the proliferation of donor-derived alloreactive T-cells, thus protecting from aGVHD (23). Although these findings are not directly translatable to human aGVHD due to specificities of HSCT models in mice and differences between mouse and human MAIT cells (23, 48, 49), they support our observations that human MAIT cells do not contribute to aGVHD. Using a classical *in vitro* model of alloreactivity, we show that MAIT cells do not proliferate in response to allogeneic cells, unless combined MR1 ligand and cytokine signals are present. Moreover, MAIT cells do not participate in the development of xeno-GVHD in NSG mice infused with huPBMCs. It is likely that the xeno-GVHD is caused by a fraction of T cells having a low frequency in donor PBMCs, which subsequently expand in mouse organs upon recognition of murine MHC (50). One cannot exclude that the NSG host with an ablated immune compartment may be less likely to provide conditions for presentation of MR1-ligands to MAIT cells or the delivery of costimulatory signals. However, MR1 is highly conserved across various species, with 90% of sequence similarity between mice and humans, so that murine MR1 can present 5-OP-RU to human MAIT cells as efficiently as human MR1 (35). That donor T cells may outcompete MAIT cells by limiting the availability of IL-15 seems unlikely, as demonstrated by the failure of exogenous human IL-15 to allow MAIT cell accumulation in tissues at time of xeno-GVHD. Whether MAIT cells fail to traffic or find their niche in the host due to species barriers between chemokine receptors and their ligands is also unlikely since MAIT cells were found in the spleen and liver one week after transfer. Rather, it appears that MAIT cells do not expand upon recognition of murine MHC and consequently do not participate to GVHD tissue lesions.

Due to ubiquitous expression of MR1 and abundance of human MAIT cells in tissues, MAIT cell functions need to be tightly regulated to control the balance between healthy and pathological processes. Following HSCT, homeostatic cytokines may provide early but limited proliferation signals to graft-derived MAIT cells, at least ensuring their survival in the recipient in the early post-transplant period. However, sustained expansion of mature and/or thymus-derived naïve MAIT cells will only occur if MR1 ligands are present together with inflammatory signals. Our data showing the lack of alloreactive potential of MAIT c ells pave the way for safely harnessing their functions in adoptive immune therapy. MAIT cells are excellent candidates for engineering universal chimeric antigen receptor cells (CAR-MAIT), as they are easy to expand *in vitro*, can traffic to tissues, have potent effector functions and are unlikely to cause GVHD. In addition, the novel tissue repair and regulatory functions of MAIT cells open up new avenues for exploiting these cells for clinical benefit.

## Data Availability

N/A

## Acknowledgments

The authors thank all the patients and their physicians and the nurse and technician staff from Hôpital Robert Debré who helped with this study, as well as all members of the CRYOSTEM Consortium for providing patients samples used in this study : University Hospital of Angers, University Hospital of Dijon Bourgogne, University Hospital of Besançon, University Hospital of Grenoble, University Hospital of Lille, University Hospital of Lyon, University Hospital of Bordeaux, Paoli-Calmettes Institute, AP-HM, University Hospital of Nantes, AP-HP (Saint Louis Hospital, La Pitié-Salpêtrière Hospital, Saint-Antoine Hospital, Robert Debré Hospital, Necker Hospital, Groupe Hospitalier Henri Mondor), INSERM, University Hospital of Toulouse, University Hospital of Tours, University Hospital of Rennes, University Hospital of Clermont-Ferrand, University Hospital of Saint-Etienne, Lucien Neuwirth Cancer Institute, University Hospital of Poitiers, University Hospital of Nice, University Hospital of Brest, Military Hospital of Percy, University Hospital of Montpellier, Gustave Roussy Institute, University Hospital of Limoges, University Hospital of Caen, Etablissement Français du Sang. CRYOSTEM project has been funded by ANR, the Institut National du Cancer (INCa) and some patient associations.

The authors are grateful to Véronique Parietti (Plateforme d’expérimentation animale, IRSL, Hôpital Saint-Louis) for her help.

## Abbreviations

CFSE: Carboxyfluorescein succinimidyl ester
GVHD: graft-versus-host disease
GVL: graft-versus-leukemia
HSCT: hematopoietic stem cell transplantation
MAIT: Mucosal associated invariant T
MA: myeloablative
NMA: non myeloablative
MSD: matched sibling donor
MUD: matched unrelated donor
TCR: T cell receptor
Tconv: conventional T cells
5-OP-RU: 5-(2-oxopropylideneamino)-6-D-ribitylaminouracil

