## supplemental methods for "Human MAIT cells are devoid of alloreactive potential: prompting their use as universal cells for adoptive immune therapy"

### Supplementary Methods

*Flow cytometry*

Multiparametric 14-color flow cytometry analyses were performed using combinations of the following antibodies for 20 min at 4°C: anti-CD3 Percp Vio770 or APC Vio770 (Miltenyi Biotec); anti-TCR V$\alpha$7.2 BV605 (clone 3C10), anti-CD161 APC-Cy7, anti-CD4 AF700 or BV711, anti-CD25 BV421, anti-CD69 APC, anti-CD45 AF700 (all from Biolegend); anti-CD8 HV500 (BD Biosciences); CD45RO FITC (Beckman Coulter).
PE-conjugated MR1:5-OP-RU or MR1:6-FP control tetramer (dilution 1:600) was performed for 45 min at room temperature before surface staining with other antibodies for 20 min at 4°C. Dead cells were excluded using Viaprobe (BD Biosciences). For intracellular staining, cells were first stained with antibodies against surface antigens, then fixed/permeabilized, washed and incubated with anti-Ki67 BV711 (Miltenyi Biotec) at 4°C for 20 minutes. Data were acquired on a BD LSR Fortessa or FACSCelesta flow cytometer (BD Biosciences). A total of at least 100,000 events in a live gate were collected. The gating strategy was CD45 versus side scatter. Gates were defined through isotype and fluorescence minus one (FMO) stains. MAIT frequencies were expressed as a percentage of CD3+ lymphocytes. Absolute numbers (per microliter) were calculated from the absolute lymphocyte count determined on the same sample with a hematology automated analyzer. Data were analyzed using FlowJo software.

*Isolation of cells from mouse tissues*

Briefly, blood, spleen and liver were prepared as single-cell suspensions. Lungs and colon were placed in RPMI medium and digested with collagenase I (lungs) or collagenase VIII (colon) and DNAse I. PBMCs from liver, lungs and colon were isolated by Percoll density gradient centrifugation. Red blood cells from the blood and spleen were lysed using FACS lysis solution (BD Biosciences) and ACK solution (Thermo Fisher), respectively.
